# The COVID-19 pandemic impact on continuity of care provision on rare brain diseases and on Ataxia, Dystonia and PKU. A scoping review protocol

**DOI:** 10.1101/2022.07.26.22277799

**Authors:** Sara Cannizzo, Vinciane Quoidbach, Paola Giunti, Wolfgang Oertel, Gregory Pastores, Maja Relja, Giuseppe Turchetti

**Affiliations:** Sant’Anna School of Advanced Studies, Institute of Management, Pisa, Italy; European Brain Council, Brussels, Belgium; University College London Hospital, United Kingdom; Philipps-Universitat Marburg, Germany; National Centre for inherited Metabolic Disorders, Mater Misericordiae University Hospital, Dublin, Ireland; University of Zagreb Medical School, Croatia

## Abstract

One of the most relevant challenges for healthcare providers during the COVID – 19 pandemic has been assuring the continuity of care to patients with complex health needs such as people living with rare diseases (RDs). The COVID–19 pandemic accelerated the healthcare sector’s digital transformation agenda. The delivery of telemedicine services instead of many face-to-face procedures has been expanded and, many healthcare services not directly related to COVID-19 treatments shifted online remotely. Many hospitals, specialist centres, patients and families started to use telemedicine because they were forced to. This trend could directly represent a good practice on how care services could be organized and continuity of care could be ensured for patients. If done properly, it could boast improved patient outcomes and become a post COVID-19 major shift in the care paradigm. There is a fragmented stakeholders spectrum, as many questions arise on: how is e-health interacting with ‘traditional’ healthcare providers; about the role of the European Reference Networks (ERNs); if can remote care retain a human touch and stay patient centric. Here, we outline a protocol for a scoping review that investigates this topic, focusing on continuity of care and novel methods (e.g., digital approaches) used to reduce the care disruption. This scoping review protocol was designed according to the Preferred Reporting Items for Systematic Reviews and Meta-Analyses extension for scoping reviews (PRISMA-ScR) standards and will culminate in a narrative synthesis of evidence.

## Introduction

### Rationale

The COVID-19 disease is an infectious disease caused by the new coronavirus that was first identified at the beginning of 2020 in Wuhan, China. The World Health Organization (WHO) declared the COVID-19 a global pandemic on 11 March 2020. Globally, there have been 521.920.560 confirmed cases of COVID-19, including 6.274.323 deaths, reported to WHO. In Europe there have been 219.393.358 confirmed cases of COVID-19 [1]. Due to the unexpected outbreak of the COVID-19, healthcare systems needed to reorganise the systems concentrating the resources for the care of COVID-19 patients and responding to this health emergency. During the emergency, the discontinuation, the interruption or delay of care, the reorganization of the patients’ care, the respect of the social/physical distancing rules had been some of the main challenges managed by the healthcare organizations including specialist centres/centres of expertise. During the emergency, many rare disease (RDs) patients experienced a very limited access to the diagnostic process and consequently they did not complete their diagnosis pathway; they experienced the interruption of their monitoring care due to the suspension of outpatient and inpatient clinics; they did not access to their healthcare providers because of the impossibility of performing face-to-face evaluations, etc. [2]

Telemedicine and telehealth were often proposed as alternative tools to face-face consultations and offered innovative solutions for managing the provision of healthcare services in emergency situations [3] [4]. Furthermore, this scoping review aims to synthesize data pertaining to strategies that have been implemented for rare neurological diseases and neurometabolic disorders care during this period, as well as the impacts of these strategies. It is expected that these strategies will consist predominantly of digital or electronic health tools, which even prior to the pandemic were being introduced in attempts to enhance care and treatment across the health spectrum.

### Objectives

The main aim of this scoping review is to investigate the impact of COVID-19 pandemic on the continuity of care provision on rare neurological diseases and neurometabolic disorders, and, in particular, on Ataxia, Dystonia and Phenylketonuria (PKU) in Europe. The specific research questions were formulated and defined by consensus among authors using the PCC (Population–Concept–Context) mnemonic [5]. The two research questions are:

1. What have been the changes of the provision of healthcare services for the care of people living with rare neurological diseases and, in particular, with Ataxia, Dystonia and Phenylketonuria (PKU) in Europe during COVID-19 pandemic?
2. What measures (like telemedicine and digital e-tools, etc.) have been implemented for the mitigation of disruption or discontinuity of care for people with RNDs in Europe during COVID-19 pandemic?

**Table 1.**
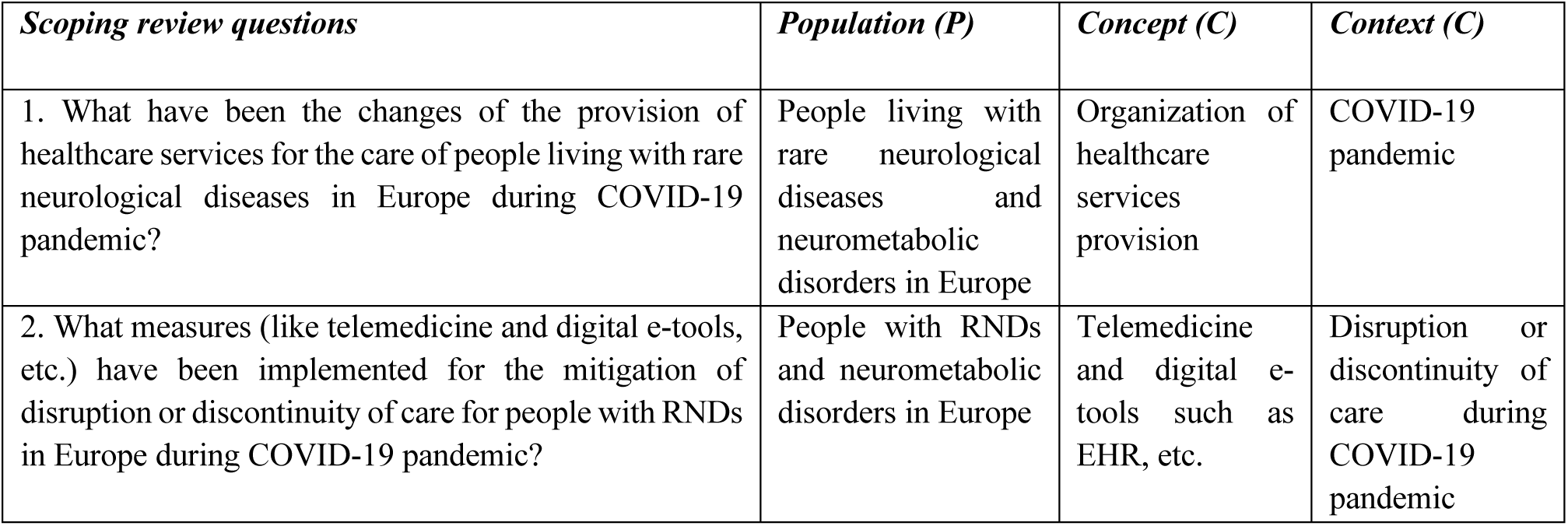
Scoping review questions

## Methods

### Design

The scoping review will be conducted in accordance with the Preferred Reporting Items for Systematic Reviews and Meta-Analyses extension for scoping reviews (PRISMA-ScR) statement [6]. The PRISMAScR statement was developed to standardise the greatly heterogenous nature of current scoping review methodology, and comprises a checklist of 22 reporting items, which was developed by an expert panel following recommendations from the Enhancing the QUAlity and Transparency Of health Research (EQUATOR) Network.

The tasks of this review are as follows: (1) defining the key review questions and scope of review, (2) searching and screening studies that meet eligibility criteria based on title and abstracts, (3) identifying relevant studies by screening full texts, (4) extracting and evaluating data, (5) constructing a narrative synthesis summarizing the results.

### Eligibility criteria

To be included in the review the articles should contain a specific field regarding the continuity of care for patients with complex health needs such as people living with rare neurological diseases (RNDs) during the COVID-19 pandemic. In particular, the changes of the provision of healthcare services for the care of people living with Ataxia, Dystonia and PKU in Europe, and the measures (like telemedicine and digital e-tools, etc.) that have been implemented for the mitigation of disruption or discontinuity of care. Moreover, we will include all publications examining any dimension of telemedicine such as the synchronous telehealth between health care providers and the patients and families, the monitoring via apps, smart devices, etc.

Peer-review articles will be included if:

a) Studies collected data during the COVID-19 pandemic, from January 2020 to the date of our search.
b) Studies conducted in Europe, written in English, and involving human participants.
c) Papers will be excluded if they will not fit into the conceptual framework of the study, focused on the continuity of care, care pathways, strategies for maintaining provision of care, improving the care provision, and novel methods used to reduce the care disruption in the field of rare neurological diseases, and in particular Ataxia, Dystonia and Phenylketonuria.
d) Observational or interventional studies were conducted, relevant systematic reviews (with or without meta-analysis), relevant scoping reviews.
e) Editorial articles, pre-print articles, opinion articles and preclinical studies will be excluded.

### Information sources and search strategy

Four electronic databases will be interrogated: PUBMED, Embase, Scopus and Web of Science from the 1^st^ of January 2020 to the search date, using search strings relevant for COVID-19 pandemic, rare neurological diseases and rare neurometabolic disorders, Ataxia, Dystonia and PKU. The search will be conducted by the first author and will be peer reviewed within the research team.

### The search strategy

((COVID-19 or COVID 2019 or severe acute respiratory syndrome coronavirus 2 or 2019 nCoV or SARSCOV2 or 2019nCoV or novel coronavirus) AND (‘rare diseases’ or ‘rare neurological diseases’ or ‘rare neurometabolic disorders’ or Ataxia or Dystonia or PKU or Phenylketonuria) AND (‘care pathway’ or organization or ‘integrated care’ ‘seamless care’ or ‘specialist centres’ or ‘centres of expertise’ or telemedicine or ‘digital health tools’ or continuity or ‘patient care pathway’ or ‘delivery of care’ or economics))

### Methodological quality assessment

Since this is a scoping review, we will not formally rate the methodological quality of included studies.

### Screening and data extraction

The search results will be exported in Mendeley, and any duplicates will be removed. Two researchers will independently screen titles and abstracts using the literature search results to select papers that meet the inclusion criteria. If necessary, disagreements will be managed to reach a consensus. If needed a third reviewer will be involved. The selected studies will move to the further step and the full texts will be reviewed. The two researchers will independently review the full study text and select the papers to be included in the final review.

### Strategy for data synthesis

A data collection sheet will be developed by the research team to extract studies characteristics including citation details (title, author, journal, date, volume, issue, pages), peer-reviewed or grey literature, country/countries involved, aims of the study, methodology, study participant (disease, age, sex, etc.), type of care setting, type of healthcare service provided, the health professionals involved in the treatment(s), the technologies used, key findings and recommendations from the studies. The results of the selected studies will be summarized by means of tables. Moreover, figures and graphs may be presented to illustrate linked themes that arise from the findings of the scoping review. A narrative synthesis will be presented to explore patterns and themes that arise from the included studies.

## Data Availability

All data produced in the present study are available upon reasonable request to the authors

